# Evaluating the Impact of Socioeconomic Factors on Dietary Choices and Nutritional Status of Adults in Selected Urban and Rural Communities in Ekiti State

**DOI:** 10.64898/2026.03.31.26349759

**Authors:** Bosede Mary Adeniyi, Abolaji Moses Ogunetimoju, Olajumoke Aina Olofinsanmabo

## Abstract

**Introduction:** Adults in Nigeria face a growing nutrition challenge: while some struggle with undernutrition, others are increasingly affected by overweight and obesity. This “double burden of malnutrition” reflects socioeconomic divides, where income, education, and household conditions shape food choices and health. This study examined how socioeconomic factors influence dietary diversity, nutritional knowledge, and nutritional status among adults in urban and rural communities of Ekiti State.

**Methods and Analysis:** A descriptive cross-sectional survey was conducted among 350 adults selected via multistage sampling. Data were collected using structured questionnaires and anthropometric measurements. Dietary diversity was assessed using food group frequency, and nutritional status was determined by BMI. Associations were analyzed using chi-square tests (p < 0.05).

**Results:** Significant urban-rural divides were identified: urban respondents were more educated (48.8% tertiary), while rural households were more dependent on farming (35.0%) and low-income (62.0% < 70,000). Dietary patterns differed significantly: urban diets favored legumes (58.4%) and cereals (56.0%), while rural diets predominated in sugar/honey (90.0%) and roots/tubers (71.0%). Nutritional knowledge was higher in urban areas. Nutritional status revealed a dual burden: 20.4% of urban and 22.0% of rural respondents were underweight, while combined overweight/obesity affected 18.4% of urban and 25.0% of rural participants.

**Conclusion:** Socioeconomic factors strongly dictate dietary choices and health in Ekiti State. Urban areas show greater diversity but rising obesity risks, while rural areas face persistent undernutrition. These findings highlight the need for tailored, state-specific interventions addressing both food insecurity and emerging diet-related chronic disease risks.

**What is already known on this topic:** Socioeconomic status strongly shapes dietary choices and nutrition in Nigeria, with urban–rural disparities and uneven nutrition knowledge contributing to both undernutrition and rising obesity, yet localized evidence among adults in Ekiti State remains scarce.

**What this study adds:** This study provides the first localized evidence from Ekiti State showing how socioeconomic disparities drive distinct urban–rural dietary patterns, confirm the coexistence of underweight and overweight adults, reveal limits of nutrition knowledge in shaping diets, and highlight the urgent need for tailored interventions that address both rural undernutrition and urban obesity.

**How this study might affect research, practice or policy:** This study shines a light on how everyday realities—income, education, and household size—shape what adults eat in Ekiti State. For research, it fills a gap by focusing on adults rather than children and offers a framework that others can use in different Nigerian states. For practice, it shows that nutrition education alone isn’t enough in cities, where affordability and access matter more, while in rural areas knowledge makes a bigger difference. For policy, it confirms that undernutrition and obesity coexist, urging leaders to move beyond one-size-fits-all national strategies and design state-specific programs that tackle food insecurity in rural areas and rising obesity in urban centers.

## 1.0 INTRODUCTION

Globally, dietary patterns and nutritional status have become central to public health due to their strong association with the rising burden of both undernutrition and non-communicable diseases (NCDs). Unhealthy diets are now recognized as one of the leading risk factors for mortality worldwide, contributing significantly to conditions such as cardiovascular diseases, diabetes, and obesity (GBD 2019 Risk Factors Collaborators, 2020). Increasingly, evidence shows that dietary behaviors are not merely individual choices but are shaped by broader socioeconomic determinants, including income, education, occupation, and living environment (Darmon & Drewnowski, 2015; Afshin et al., 2019). These factors influence access to food, dietary quality, and nutritional outcomes, thereby reinforcing health inequalities across populations.

In sub-Saharan Africa, the situation is further complicated by rapid urbanization and ongoing nutrition transition. Many countries in the region are experiencing a shift from traditional diets rich in whole foods to more processed, energy-dense diets, particularly in urban areas (Popkin et al., 2020). At the same time, rural populations continue to face structural constraints such as poverty, limited food access, and seasonal food insecurity, resulting in persistent undernutrition (FAO et al., 2023). This dual dynamic has led to the emergence of the “double burden of malnutrition,” where undernutrition coexists with overweight, obesity, and diet-related NCDs within the same populations (WHO, 2021).

Nigeria reflects these broader regional trends, with significant disparities in dietary patterns and nutritional outcomes across socioeconomic and geographic groups. While urban populations often have greater access to diverse and market-based food systems, they are also more exposed to processed and unhealthy food options. Conversely, rural populations frequently depend on subsistence agriculture and limited food markets, which can restrict dietary diversity and nutrient intake (National Bureau of Statistics [NBS], 2022). Socioeconomic inequalities, including income disparities, educational attainment, and occupational differences, further shape these dietary behaviors and contribute to uneven nutritional outcomes across the country (Olatona et al., 2018; Amugsi et al., 2017). Despite growing national evidence, there remains limited context-specific research focusing on adult populations, particularly at the subnational level.

Ekiti State provides a unique context for examining these dynamics due to its mix of urban and rural settings. Ado-Ekiti, the state capital, represents a more urbanized environment with greater access to infrastructure, markets, and services, while rural communities such as Epe-Ekiti are largely agrarian with comparatively limited economic opportunities and access to resources. These structural differences are likely to influence dietary choices, food accessibility, and nutritional outcomes in distinct ways. However, empirical evidence comparing these settings remains scarce.

Against this backdrop, this study evaluates the impact of socioeconomic factors on dietary choices and nutritional status among adults in selected urban and rural communities in Ekiti State. By examining the interplay between socioeconomic characteristics, dietary patterns, and nutritional outcomes, the study aims to provide context-specific evidence to inform targeted nutrition interventions and public health strategies tailored to both urban and rural population. The aim of this study is to evaluate the impact of socioeconomic factors on dietary choices and nutritional status among adults in selected urban (Ado-Ekiti) and rural (Epe-Ekiti) communities in Ekiti State, Nigeria. Specifically, the study seeks to assess the socioeconomic characteristics of the study population, examine household dietary diversity and food consumption patterns, evaluate nutritional knowledge, and determine the nutritional status of adults in both settings. In addition, the study aims to analyze the relationships between socioeconomic variables, dietary practices, and nutritional outcomes in order to generate evidence for context-specific public health interventions. By enhancing access to nutrition education and supporting food security initiatives, policymakers can significantly improve health outcomes and combat malnutrition, especially in vulnerable populations. The collective insights from these studies underscore the importance of comprehensive strategies aimed at fostering healthier eating practices across different segments of society in Nigeria.

## 2.0 METHODOLOGY

This study adopted a structured and systematic approach to examine the relationship between socioeconomic factors, dietary choices, and nutritional status among adults in urban and rural Ekiti State.

### 2.1 Study Design

A descriptive cross-sectional survey design was employed to provide a snapshot of the study population at a single point in time. This design was appropriate for assessing patterns, relationships, and variations in dietary behavior and nutritional outcomes across different socioeconomic groups.

### 2.2 Study Area and Population

The study was conducted in Ado-Ekiti (urban) and Epe-Ekiti (rural) in Ekiti State, Nigeria. The target population comprised adults aged ≥20 years who had resided in the study areas for at least one year. The combined population of both areas was estimated at approximately 658,201.

### 2.3 Sample Size Determination

The sample size was determined using Cochran’s formula for proportions:

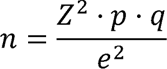

Where:

- n= required sample size
- z= standard normal deviation (1.96 at 95% confidence level)
- p= estimated proportion (0.683)
- *q = 1 - p*
- e= margin of error (0.05)

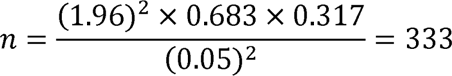

Adjusting for a 5% non-response rate yielded a final sample size of 350 participants.

### 2.4 Sampling Technique

A multistage sampling technique was used to ensure representativeness:

- **Stage 1:** Random selection of two wards per community
- **Stage 2:** Systematic sampling of households within selected wards
- **Stage 3:** Random selection of one eligible adult per household

### 2.5 Data Collection Instrument

Data were collected using a structured questionnaire covering:

- Socio-demographic characteristics
- Socioeconomic status
- Dietary patterns and food consumption
- Nutritional knowledge and perceived health status

The instrument included mainly closed-ended questions to ensure consistency and ease of analysis.

### 2.6 Data Collection Procedure

Data were collected through face-to-face interviews conducted by trained assistants. Questionnaires were administered in English or Yoruba to ensure comprehension. Daily checks were performed to ensure completeness and accuracy.

### 2.7 Validity and Reliability

Content and face validity were established through expert review. Reliability was assessed using a test–retest method among 20 participants, yielding a correlation coefficient of 0.74, indicating acceptable reliability.

### 2.8 Study Variables

- Independent variables: Age, gender, education, occupation, income, and place of residence
- Dependent variables: Dietary choices, dietary diversity, and nutritional status

### 2.9 Data Analysis

Data were analyzed using SPSS. Descriptive statistics (means, frequencies, standard deviations) were used to summarize variables, while inferential statistics (Chi-square) was applied to test associations at a significance level of p < 0.05.

### 2.10. Patient and Public Involvement Statement

Patients or members of the public were not involved in the design, conduct, reporting, or dissemination plans of this research. The study focused on evaluating the impact of socioeconomic factors on dietary choices and nutritional status among adults in selected urban and rural communities in Ekiti State. Research participants provided primary data through structured surveys and nutritional assessments; however, they did not serve as active collaborators in the data analysis or manuscript preparation. The research questions and outcome measures were developed based on gaps identified in existing nutritional literature. Findings will be disseminated to the relevant health authorities in Ekiti State and shared with the participating communities through appropriate institutional channels at Afe Babalola University.

### 2.11 Ethical Considerations

Ethical approval was obtained from the Afe Babalola University Ethics Committee. Informed consent was obtained from all participants, and confidentiality and anonymity were strictly maintained.

## 3.0 RESULTS

The results in Table 1 reveal a clear urban–rural divide in both socioeconomic status and living conditions. Urban respondents in Ado-Ekiti were generally younger, more educated, and economically advantaged, with nearly half having tertiary education (48.8%) and a more balanced income distribution. In contrast, Epe-Ekiti showed a higher concentration of older adults, lower educational attainment, and a significantly larger proportion of low-income earners (62.0% earning < 70,000), highlighting structural economic disadvantage. Occupational patterns further reinforce this divide: Ado-Ekiti is dominated by formal and service-based jobs, while Epe-Ekiti is largely agrarian, with farming as the primary occupation (35.0%). This directly influences food access, as urban residents rely almost entirely on markets (96.8%), whereas rural residents depend heavily on subsistence farming (51.0%). Household structure also differs, with larger household sizes more common in rural areas (43.0%), suggesting increased pressure on limited resources. Additionally, living conditions and infrastructure are markedly unequal, as urban residents predominantly live in cement housing (100%) with better sanitation (70.0% water closet), while rural residents experience poorer housing and high rates of open defecation (46.0%).

Overall, these findings indicate that socioeconomic inequality, occupation, and infrastructure gaps are key drivers shaping dietary access and nutritional vulnerability between urban and rural populations.

**Table 4.1:** Socio-Demographic and Economic Characteristics of Respondents (n = 350).

| Variable | Categories | Ado-Ekiti (n=250) | % | Epe-Ekiti (n=100) | % |
| --- | --- | --- | --- | --- | --- |
| Age (years) | 20–29 | 65 | 26.0 | 16 | 16.0 |
|  | **30–39** | **48** | **19.2** | **18** | **18.0** |
|  | **40–49** | **40** | **16.0** | **12** | **12.0** |
|  | **50–59** | **51** | **20.4** | **39** | **39.0** |
|  | **≥60** | **46** | **18.4** | **15** | **15.0** |
| **Mean ± SD** |  | **43.6 ± 14.67** |  | **46.0 ± 13.32** |  |
| **Sex** | **Male** | **133** | **53.2** | **51** | **51.0** |
|  | **Female** | **117** | **46.8** | **49** | **49.0** |
| **Marital Status** | **Married** | **153** | **61.2** | **14** | **14.0** |
|  | **Single** | **72** | **28.8** | **47** | **47.0** |
|  | **Divorced/Widowed** | **25** | **10.0** | **39** | **39.0** |
| **Education** | **None** | **30** | **12.0** | **29** | **29.0** |
|  | **Primary** | **23** | **9.2** | **23** | **23.0** |
|  | **Secondary** | **22** | **8.8** | **10** | **10.0** |
|  | **Tertiary** | **122** | **48.8** | **24** | **24.0** |
|  | **Others** | **53** | **21.2** | **14** | **14.0** |
| **Occupation** | **Farmer** | **43** | **17.2** | **35** | **35.0** |
|  | **Artisan** | **38** | **15.2** | **22** | **22.0** |
|  | **Self-employed** | **25** | **10.0** | **15** | **15.0** |
|  | **Civil servant** | **50** | **20.0** | **6** | **6.0** |
|  | **Teacher** | **53** | **21.2** | **12** | **12.0** |
|  | **Professional** | **41** | **16.4** | **10** | **10.0** |
| **Household Size** | **<3** | **76** | **30.4** | **30** | **30.0** |
|  | **3–4** | **104** | **41.6** | **27** | **27.0** |
|  | **≥5** | **70** | **28.0** | **43** | **43.0** |
| **Monthly Income (□)** | **<70,000** | **71** | **28.4** | **62** | **62.0** |
|  | **70,000–120,000** | **82** | **32.8** | **6** | **6.0** |
|  | **120,000–170,000** | **47** | **18.8** | **18** | **18.0** |
|  | **≥170,000** | **50** | **20.0** | **14** | **14.0** |
| **Main Food Source** | **Farming** | **6** | **2.4** | **51** | **51.0** |
|  | **Market** | **242** | **96.8** | **27** | **27.0** |
|  | **Gifts** | **2** | **0.8** | **10** | **10.0** |
|  | **Food aid** | **–** | **–** | **12** | **12.0** |
| **Housing Type** | **Mud (thatch)** | **–** | **–** | **12** | **12.0** |
|  | **Mud (zinc)** | **–** | **–** | **28** | **28.0** |
|  | **Cement** | **250** | **100** | **60** | **60.0** |
| **Cooking Fuel** | **Firewood** | **36** | **14.4** | **30** | **30.0** |
|  | **Charcoal** | **93** | **37.2** | **12** | **12.0** |
|  | **Kerosene** | **18** | **7.2** | **7** | **7.0** |
|  | **Gas** | **80** | **32.0** | **26** | **26.0** |
|  | **Electricity** | **23** | **9.2** | **25** | **25.0** |
| **Water Source** | **Pipe-borne** | **43** | **19.2** | **25** | **25.0** |
|  | **Borehole** | **86** | **34.4** | **6** | **6.0** |
|  | **Well** | **56** | **22.4** | **48** | **48.0** |
|  | **Stream** | **18** | **7.2** | **11** | **11.0** |
|  | **Rainwater** | **12** | **4.8** | **6** | **6.0** |
|  | **Sachet water** | **35** | **14.0** | **4** | **4.0** |
| **Toilet Facility** | **Water closet** | **175** | **70.0** | **22** | **22.0** |
|  | **Pit latrine** | **65** | **26.0** | **28** | **28.0** |
|  | **Bucket** | **–** | **–** | **2** | **2.0** |
|  | **Open defecation** | **10** | **4.0** | **46** | **46.0** |

**Source:** Field survey,2025

Table 4.2 shows the Household Dietary Diversity among the 350 respondents from Ado-Ekiti and Epe-Ekiti. The data reveals distinct patterns in food consumption between the two locations. In Ado-Ekiti, the most commonly consumed food groups are Legumes 146(58.4%), Cereals 140(56.0%), Vegetables 140(56.0%), and Fruits 142(56.8%). Conversely, the least consumed food groups in Ado-Ekiti are Nuts & Seeds 101(40.4%), Sugar or Honey 109(43.6%), and Fish & Fish Products 113(45.2%). In contrast, the dietary pattern in Epe-Ekiti is markedly different. The consumption of Sugar or Honey is exceptionally high (90.0%), followed by Fruits (78.0%) and Roots & Tubers (71.0%). Epe-Ekiti also shows a notably higher consumption of Nuts & Seeds (61.0%) compared to Ado-Ekiti. The food groups with the lowest consumption in Epe-Ekiti are Milk & Dairy (35.0%) and Cereals (43.0%).

**Table 4.2:** Household Dietary Diversity (n = 350).

| Food Group | Examples | Ado-Ekiti Yes (%) | Ado-Ekiti No (%) | Epe-Ekiti Yes (%) | Epe-Ekiti No (%) |
| --- | --- | --- | --- | --- | --- |
| **Cereals** | Bread, rice, sorghum, millet, wheat, maize, biscuits, etc. | 140 (56.0%) | 110 (63.2%) | 43 (43.0%) | 57 (57.0%) |
| **Root & Tuber** | Yam, potatoes, cassava, etc. | 128 (51.2%) | 122(61.2%) | 71 (71.0%) | 29 (29.0%) |
| **Vegetables** | Pumpkin, spinach, pepper, tomatoes | 140 (56.0%) | 110 (44.0%) | 47 (47.0%) | 53 (53.0%) |
| **Fruits** | Orange, banana, pawpaw, apple, etc. | 142 (56.8%) | 108(43.2%) | 78 (78.0%) | 22 (22.0%) |
| **Meat** | Beef, chicken, goat, pork | 135 (54.0%) | 115 (46.0%) | 47 (47.0%) | 53 (53.0%) |
| **Eggs** | Eggs | 129 (51.6%) | 121(48.4%) | 54 (54.0%) | 46 (46.0%) |
| **Fish & Fish Products** | Sardines, dried fish, catfish, crayfish | 113 (45.2%) | 137(54.8%) | 47 (47.0%) | 53 (53.0%) |
| **Legumes** | Beans, soybean, melon, sesame, locust bean | 146 (58.4%) | 104(41.6%) | 45 (45.0%) | 55 (55.0%) |
| **Milk & Dairy** | Milk, yoghurt, cheese, wara | 126 (50.4%) | 124(49.6%) | 35 (35.0%) | 65 (65.0%) |
| **Nuts & Seeds** | Groundnut, coconut, cashew, walnut | 101 (40.4%) | 149(59.6%) | 61(67.0%) | 39 (39.0%) |
| **Oil, Fat & Butter** | Cooking oil, butter, margarine | 116 (46.4%) | 134(53.6%) | 51 (51.0%) | 49 (49.0%) |
| **Sugar or Honey** | Sugar, sweetened drinks, honey, chocolate | 109 (43.6%) | 141(56.4%) | 62 (90.0%) | 38 (38.0%) |

**Source:** Field survey,2025

Table 4.3, reveals significant differences in how often various food groups are consumed between the two locations. A prominent trend shows that respondents in Epe-Ekiti are the most frequent consumers of “Yam& Yam Products,” with nearly half the population (47.0%, n=47) eating them five or more times a week, a rate substantially higher than in Ado-Ekiti (16.0%, n=40).

**Table 4.3:** Food frequency of the respondents (n = 350).

| **Food Group** | **Location** | **Never** | **1–2x/week** | **3–4x/week** | **≥5x/week** |
| --- | --- | --- | --- | --- | --- |
| **Yam & Yam Products** | Ado-Ekiti | 68 (27.2%) | 65 (26.0%) | 77 (30.8%) | 40 (16.0%) |
|  | Epe-Ekiti | 19 (19.0%) | 8 (8.0%) | 26 (26.0%) | 47 (47.0%) |
| **Potatoes** | Ado-Ekiti | 50 (20.0%) | 48 (19.2%) | 82 (32.8%) | 70 (28.0%) |
|  | Epe-Ekiti | 30 (30.0%) | 29 (29.0%) | 37 (37.0%) | 4 (4.0%) |
| **Cassava Products** | Ado-Ekiti | 56 (22.4%) | 74 (29.6%) | 58 (23.2%) | 62 (24.8%) |
|  | Epe-Ekiti | 32 (24.0%) | 19 (19.0%) | 43 (43.0%) | 6 (6.0%) |
| **Cocoyam** | Ado-Ekiti | 77 (30.8%) | 59 (23.6%) | 53 (21.2%) | 61 (24.4%) |
|  | Epe-Ekiti | 22 (22.0%) | 30 (30.0%) | 20 (20.0%) | 28 (28.0%) |
| **Plantain** | Ado-Ekiti | 47 (18.8%) | 97 (38.8%) | 42 (16.8%) | 64 (25.6%) |
|  | Epe-Ekiti | 20 (20.0%) | 37 (37.0%) | 27 (16.0%) | 16(16.0%) |
| **Rice** | Ado-Ekiti | 50 (20.0%) | 46 (18.4%) | 71 (28.4%) | 83 (33.2%) |
|  | Epe-Ekiti | 26 (26.0%) | 22 (22.0%) | 28 (28.0%) | 24 (24.0%) |
| **Wheat** | Ado-Ekiti | 60 (24.0%) | 58 (23.2%) | 52 (20.8%) | 80 (32.0%) |
|  | Epe-Ekiti | 43 (43.0%) | 29 (29.0%) | 18 (18.0%) | 10 (10.0%) |
| **Oats** | Ado-Ekiti | 54 (21.6%) | 50 (20.0%) | 69 (27.6%) | 77 (30.8%) |
|  | Epe-Ekiti | 16 (16.0%) | 35 (35.0%) | 18 (18.0%) | 31 (25.0%) |
| **Beans & Bean Products** | Ado-Ekiti | 63 (25.2%) | 77 (30.8%) | 25 (10.0%) | 85 (34.0%) |
|  | Epe-Ekiti | 36 (36.0%) | 8 (8.0%) | 25 (25.0%) | 31 (31.0%) |
| **Soybeans** | Ado-Ekiti | 52 (20.8%) | 78 (31.2%) | 60 (24.0%) | 60 (24.0%) |
|  | Epe-Ekiti | 37 (37.0%) | 18 (18.0%) | 18 (18.0%) | 27 (27.0%) |
| **Groundnut** | Ado-Ekiti | 50 (20.0%) | 55 (22.0%) | 74 (29.6%) | 71 (28.4%) |
|  | Epe-Ekiti | 53 (53.0%) | 33 (33.0%) | 14 (14.0%) | - |
| **Locust Beans** | Ado-Ekiti | 31 (12.4%) | 82 (32.8%) | 86 (34.4%) | 51 (20.4%) |
|  | Epe-Ekiti | 21 (21.0%) | 25 (25.0%) | 42 (42.0%) | 12 (12.0%) |
| **Eggs** | Ado-Ekiti | 91 (36.4%) | 49 (19.6%) | 59 (23.6%) | 51 (20.4%) |
|  | Epe-Ekiti | 33 (33.0%) | 22 (22.0%) | 20 (20.0%) | 25 (25.0%) |
| **Cray Fish** | Ado-Ekiti | 57 (22.8%) | 68 (27.2%) | 82 (32.8%) | 43 (17.2%) |
|  | Epe-Ekiti | 14 (14.0%) | 44 (44.0%) | 27 (27.0%) | 30 (30.0%) |
| **Titus** | Ado-Ekiti | 75 (30.0%) | 39 (15.6%) | 54 (21.6%) | 82 (32.8%) |
|  | Epe-Ekiti | 18 (18.0%) | 32 (32.0%) | 18 (32.0%) | 32 (32.0%) |

**Source:** Field survey,2025

Conversely, for many other staples, the pattern is reversed. Consumption of Rice, Wheat, and Beans multiple times a week (≥3 times) is consistently higher in Ado-Ekiti. For instance, a third of Ado-Ekiti respondents (33.2%, n=83) eat rice five or more times a week, compared to a quarter (24.0%, n=24) in Epe-Ekiti. The disparity is even more pronounced for wheat, where 32.0% (n=80) in Ado-Ekiti are high-frequency consumers versus only 10.0% (n=10) in Epe-Ekiti.

The data also highlights specific local preferences. Cassava Products are consumed 3-4 times a week by a large portion of Epe-Ekiti respondents (43.0%, n=43), while Ado-Ekiti shows a more distributed pattern. Furthermore, a notable proportion of the population in both locations never consumes certain foods, such as Groundnut in Epe-Ekiti (53.0%, n=53 never eat it) and Eggs in Ado-Ekiti (36.4%, n=91 never eat them).

### 4.4 Anthropometric measurements of respondents (BMI)

Table 4.4, reveals notable differences in the physical characteristics of respondents between the two locations. The Body Mass Index (BMI) data provides a more standardised health indicator. It shows that the majority of respondents in both locations have a normal BMI, with a higher prevalence in Ado-Ekiti (61.2%, n=153) than in Epe-Ekiti (53.0%, n=53). The rates of overweight and obesity based on BMI are similar in both areas, with Ado-Ekiti having slightly higher combined figures (18.4%, n=46) than Epe-Ekiti (25.0%, n=25). Meanwhile, the percentage of underweight individuals is nearly identical, at 20.4% (n=51) in Ado-Ekiti and 22.0% (n=22) in Epe-Ekiti.

**Table 4.4:** Anthropometric measurements of respondents (BMI) (n=350).

| Variables | Categories | Ado-Ekiti<br>(Freq) | Ado-Ekiti<br>(%) | Epe-Ekiti<br>(Freq) | Epe-Ekiti<br>(%) |
| --- | --- | --- | --- | --- | --- |
| **Underweight** | < 18.5kg | 51 | 20.4 | 22 | 22.0 |
| **Normal weight** | 18.5 – 24.9kg | 153 | 61.2 | 53 | 53.0 |
| **Overweight** | 25.0 – 29.9kg | 25 | 10.0 | 13 | 13.0 |
| **Obese** | ≥ 30.0kg | 21 | 8.4 | 12 | 12.0 |

**Source:** Field survey,2025

Table 5 presents the association between nutritional knowledge and household dietary diversity among respondents in Ado-Ekiti and Epe-Ekiti. In Ado-Ekiti, formal nutrition education showed a significant association with dietary diversity (p = 0.031), indicating that individuals who had received nutrition education were more likely to have better dietary diversity. Similarly, knowledge of food groups (p = 0.050), awareness of nutrition-related diseases (p < 0.001), and the belief that nutrition can prevent diseases (p = 0.023) were significantly associated with dietary diversity. However, knowledge of balanced nutrition (p = 0.764) and recognition of malnutrition signs (p = 0.473) were not significantly associated.

**Table 5:** Association Between Nutritional Knowledge and Household Dietary Diversity Among Residents of Ado-Ekiti and Epe-Ekiti.

| Variables | Categories | Ado-Ekiti<br>High | Ado-Ekiti<br>Low | $\chi^2$ | P-value | Epe-Ekiti<br>High | Epe-Ekiti<br>Low | $\chi^2$ | P-value |
| --- | --- | --- | --- | --- | --- | --- | --- | --- | --- |
| Formal nutrition education | Yes | 111 | 60 | 4.668 | **0.031** | 20 | 35 | 0.625 | 0.429 |
|  | No | 62 | 17 |  |  | 13 | 32 |  |  |
| Knowledge of balanced diet | Yes | 89 | 38 | 0.539 | 0.764 | 19 | 42 | 21.836 | **<0.001** |
|  | No | 42 | 17 |  |  | 3 | 16 |  |  |
|  | Not sure | 42 | 22 |  |  | 14 | 6 |  |  |
| **Knowledge of food groups** | Carbohydrates | 12 | 5 | 9.047 | **0.050** | 11 | 11 | 14.130 | **0.001** |
|  | Proteins | 9 | 1 |  |  | - | - |  |  |
|  | Fats & oils | 2 | 4 |  |  | 14 | 13 |  |  |
|  | Vitamins | 2 | 0 |  |  | - | - |  |  |
|  | All food groups | 147 | 68 |  |  | 14 | 13 |  |  |
| **Signs of malnutrition** | Underweight | 3 | 0 | 1.499 | 0.473 | 11 | 13 | 2.495 | 0.287 |
|  | Overweight | 6 | 2 |  |  | 4 | 12 |  |  |
|  | All of the above | 164 | 75 |  |  | 18 | 42 |  |  |
| **Diseases linked to poor nutrition** | Malaria | 30 | 17 | 7.384 | **<0.001** | 11 | 27 | 7.384 | 0.054 |
|  | Diabetes | 41 | 5 |  |  | 4 | 12 |  |  |
|  | HIV | 55 | 17 |  |  | 14 | 12 |  |  |
|  | Typhoid | 47 | 38 |  |  | 4 | 16 |  |  |
| **Nutrition prevents disease** | Yes | 101 | 31 | 7.526 | **0.023** | 23 | 42 | 15.823 | **<0.001** |
|  | No | 33 | 24 |  |  | 2 | 17 |  |  |
|  | Not sure | 39 | 22 |  |  | 10 | 6 |  |  |

**Source:** Field survey,2025

In Epe-Ekiti, knowledge of balanced nutrition (p < 0.001), understanding of food groups (p = 0.001), and awareness that nutrition prevents diseases (p < 0.001) were significantly associated with dietary diversity. In contrast, formal nutrition education (p = 0.429), knowledge of malnutrition signs (p = 0.287), and diseases associated with poor nutrition (p = 0.054) did not show significant associations.

Overall, the findings suggest that nutritional knowledge—particularly practical understanding of balanced diets, food groups, and disease prevention—is a key determinant of dietary diversity, although the influence varies between urban and rural settings.

## 4.0 CONCLUSION

This paper presents a localized evaluation of the socioeconomic determinants shaping dietary behaviors and nutritional outcomes in Ekiti State, revealing significant urban-rural disparities and a clear structural divide between 2025 and 2026. According to the findings, the nutritional landscape in Ekiti exemplifies a “double burden of malnutrition” where undernutrition and overnutrition coexist across both settings, driven by distinct economic gradients. Urban respondents in Ado-Ekiti, characterized by higher educational attainment and market-based food access, demonstrate greater dietary diversity but face a rising prevalence of overweight and obesity. Epe-Ekiti, by contrast, remains typified by an agrarian economy and lower income levels, where dietary choices are heavily influenced by subsistence farming and a high frequency of tuber consumption, leading to persistent undernutrition.

The statistical analysis proves that these nutritional discrepancies are not accidental but are influenced by existing socioeconomic variables such as household size, occupation, and infrastructure. Although nutritional knowledge serves as a significant determinant of dietary diversity, its impact is inconsistent; in rural areas, practical understanding of balanced diets is a potent factor, whereas in urban centers, formal education and the ability to prevent disease through nutrition play more dominant roles. These findings dispute “one-size-fits-all” national nutrition strategies and demonstrate the need for decentralized, state-specific policies that effectively respond to the rising obesity risks in urban areas and food insecurity in rural communities. Policies should consequently strive to expand targeted food security initiatives and rural nutrition education while capitalizing on affordability-based interventions and lifestyle modifications in urban centers. To further strengthen evidence-based planning, future research should explore the longitudinal impact of the nutrition transition and market price fluctuations on adult health across other Nigerian subregions.

## Statements and Declarations

### Author Contributions

**Bosede Mary Adeniyi** conceptualized and designed the study, led the literature synthesis, and performed the initial drafting of the manuscript. She was responsible for defining the nutritional and socioeconomic framework used to analyze dietary disparities and managed the overall integration of the public health discussion. She supervised and led the data collection process across both urban and rural study sites. As the corresponding author, she provided final approval for the version to be published. **Abolaji Moses Ogunetimoju** designed the sampling framework, performed the formal statistical analysis using SPSS, and generated the results interpretations.

He provided critical interpretation of the data and conducted a rigorous final review and editing of the manuscript to ensure methodological consistency and intellectual quality. He also oversaw the submission process and final manuscript coordination. **Olajumoke Aina Olofinsanmabo** assisted in developing the biological and nutritional assessment framework and contributed to data interpretation. She performed the critical inline editing and revision of the manuscript for intellectual content. All authors have read and approved the final manuscript for publication

### Clinical trial number

Not applicable.

### Institutional Review Board Statement

The study was conducted in accordance with the Declaration of Helsinki and the protocol was approved by the Health Research Ethics Committee (HREC) of the Ministry of Health, Ekiti State, Nigeria. The study procedures were designed to ensure the strict confidentiality and anonymity of all participants. All subjects were informed regarding the purpose of the study, their right to voluntary participation, and their right to withdraw at any stage without penalty. Informed consent was obtained from all participants involved in the study prior to the administration of questionnaires and the collection of anthropometric measurements.

## Data Availability

The data supporting the findings of this study are available from the corresponding author upon reasonable request. The data are not publicly available due to privacy and ethical restrictions concerning participant confidentiality as outlined by the Afe Babalola University Research Ethics Committee (ABUAD-REC).

## Acknowledgments

The authors wish to express their sincere gratitude to the participants from Ado-Ekiti and Epe-Ekiti for their cooperation and willingness to share information for this study. We also acknowledge the field assistants for their dedication during the data collection process across both urban and rural communities. Special thanks to the community leaders and local authorities in the selected study areas for their invaluable support in facilitating community entry and ensuring the smooth conduct of the research. Finally, we appreciate the management and staff of the Department of Human Nutrition and Dietetics at Afe Babalola University, Ado Ekiti, and the Department of Statistics at Obafemi Awolowo University, Ile-Ife, for providing the institutional support and technical resources necessary for the completion of this research.

## REFERENCES

Afshin, A., Sur, P. J., Fay, K. A., Cornaby, L., Ferrara, G., Salama, J. S.,…& Murray, C. J. (2019). Health effects of dietary risks in 195 countries, 1990–2017: a systematic analysis for the Global Burden of Disease Study 2017. The Lancet, 393(10184), 1958–1972.

Amugsi, D. A., Mittelmark, M. B., & Kyobutungi, C. (2017). Socio-economic and demographic determinants of household food security and dietary diversity in Nigeria. Public Health Nutrition.

Darmon, N., & Drewnowski, A. (2015). Does social class predict diet quality? American Journal of Clinical Nutrition, 101(5), 908–914.

FAO, IFAD, UNICEF, WFP, & WHO. (2023). The State of Food Security and Nutrition in the World 2023: Urbanization, agrifood systems transformation and healthy diets across the rural–urban continuum. Rome: FAO.

GBD 2019 Risk Factors Collaborators. (2020). Global burden of 87 risk factors in 204 countries and territories, 1990–2019: a systematic analysis for the Global Burden of Disease Study 2019. The Lancet, 396(10258), 1223–1249.

National Bureau of Statistics (NBS). (2022). Nigeria Multidimensional Poverty Index. Abuja, Nigeria.

Olatona, F. A., Onajole, A. T., Adenihun, A. A., & Afolabi, B. B. (2018). Dietary habits and metabolic risk factors for non-communicable diseases among residents of an urban community in Lagos State, Nigeria. Journal of Community Medicine and Primary Health Care.

Popkin, B. M., Corvalan, C., & Grummer-Strawn, L. M. (2020). Dynamics of the double burden of malnutrition and the changing nutrition reality. The Lancet, 395(10217), 65–74.

World Health Organization (WHO). (2021). The double burden of malnutrition: Priority actions. Geneva: World Health Organization.

